# Multiplex serology reveals age-specific immunodynamics of endemic respiratory pathogens in the wake of the COVID-19 pandemic

**DOI:** 10.1101/2025.03.26.25324738

**Authors:** Samantha J. Bents, Emily T. Martin, Terry Steven-Ayers, Claire Andrews, Amanda Adler, Amanda Perofsky, Elizabeth M. Krantz, Rachel Blazevic, Louise Kimball, Robin Prentice, Chelsea Hansen, Lea Starita, Peter Han, Janet A. Englund, Nicole Wolter, Anne von Gottberg, Lorens Maake, Jocelyn Moyes, Cheryl Cohen, Michael Boeckh, James A Hay, Alpana Waghmare, Cécile Viboud

**Author notes:** Corresponding authors: Samantha Bents, Cecile Viboud.

## Abstract

The rebound of endemic respiratory viruses following the COVID-19 pandemic was marked by atypical transmission dynamics, with children experiencing increased disease burden and out-of-season epidemics as restrictions relaxed. Here we used serology from a newly developed quantitative multiplex assay to assess the post-pandemic immunity debt, a drop in immunity due to a lack of endemic virus circulation during COVID-19. We assessed age-specific antibody dynamics in Seattle, Washington, US, across a broad range of respiratory viruses, including influenza, respiratory syncytial virus, seasonal coronaviruses, and SARS-CoV-2. We found that respiratory virus immunodynamics differed between individuals <5 years of age and older individuals, with young children experiencing both larger boosts and quicker waning of antibodies across pathogens. We confirmed that these patterns are upheld in a non-pandemic setting by analyzing influenza serological data from South Africa. We incorporated our serological insights into an influenza transmission model calibrated to epidemiological data from Seattle and show that consideration of age-specific immunodynamics may be important to anticipate the effects of pandemic perturbations.

## Introduction

Respiratory virus epidemics are governed by a complex interplay between the waxing and waning of population immunity, cycling of different strains or subtypes, and seasonal forcing. The COVID-19 pandemic has provided a unique opportunity to clarify the forces driving epidemics. The circulation of many endemic viral pathogens was interrupted by non-pharmaceutical interventions (NPIs), only to rebound when population contacts resumed (1,2). The consequences of the so-called “immunity debt”, whereby population immunity is reduced due to lack of pathogen exposure, have been seen across various disease systems. For instance, large, out-of-season respiratory syncytial virus (RSV) outbreaks were reported globally following periods of low circulation, with several rebounds showing an age shift to older infants experiencing more severe disease (3–5). Similarly, influenza activity remained historically low for several seasons before rebounding to generally typical magnitude and timing but disrupted age structure (6,7). Pathogens such as seasonal coronaviruses, adenovirus, parainfluenza viruses, rhinovirus, and other respiratory infections experienced similar perturbations, with variable timing and extent of rebound (8,9). These pandemic perturbations allow for a rare window into how host immunity may decline in the absence of endemic virus exposure, in turn helping to clarify the contribution of immune waning to pathogen dynamics.

Serology has emerged as a powerful tool to monitor population immunity and deepen our understanding of disease dynamics (10,11). Elevated serum antibody concentration levels can be indicative of a recent pathogen disease exposure, and thus provide useful insight into the history of circulating pathogens (12,13). Age and immune function influence an individual’s antibody dynamics, where antibody concentration levels generally increase with age and exposure throughout childhood but may stabilize or decline at older ages, a phenomenon referred to as the antibody ceiling effect (14,15). In several instances, individuals have also been shown to mount a large antibody response to the first subtype of a pathogen that they are exposed to throughout life, known as antigenic seniority, and produce cross-reactive antibody responses when infected later with multiple related strains (16). Despite the complex biological processes that dictate an individual’s antibody response, population-level serology can shed light on the key mechanisms underpinning recent outbreaks and anticipate future outbreaks.

While there is a growing body of serological studies addressing repeat exposure with endemic respiratory pathogens, the persistence of immune markers in the absence of pathogen exposure remains less well-studied. The COVID-19 pandemic offers a unique opportunity to study the drivers of the decay of immune responses, particularly relating to age and prior exposures. For instance, an influenza serology study set in the pre-COVID-19 era has reported differences in the duration of immunity by age and circulating subtypes (17). Whether these age differences are common to other respiratory pathogens and whether they can be exacerbated or reduced during periods of low pathogen exposure such as the COVID-19 pandemic remains unclear. Here, we rely on a novel multiplex serological assay collected by the Seattle Flu Alliance (SFA) throughout the COVID-19 pandemic in the greater Seattle area, USA, to model the age-specific antibody dynamics of influenza, RSV, and seasonal and pandemic coronaviruses. We complement our data with pre-pandemic influenza serology collected independently in South Africa using a well-established hemagglutination inhibition assay (HAI) (18). To extend our findings across scales, we incorporate serological insights in mathematical transmission models and test hypotheses about the influenza rebound in 2022-23 observed in the United States, where children experienced the highest hospitalization rates recorded in recent seasons (19). Our study provides a unique focus on young children, generating insight into the nature of early childhood exposures and immunity to respiratory viruses, and their impact on disease dynamics.

## Results

### Modeling the age-specific immunodynamics of endemic respiratory pathogens using serological data

#### Virological and serological surveillance in King County, WA

We used virological and serological surveillance data collected as part of the Seattle Flu Alliance, a large respiratory study set in King County, WA, USA, to assess pathogen activity and antibody dynamics during the COVID-19 pandemic. In King County, respiratory virus activity was repressed throughout the early COVID-19 pandemic period (see **Fig 1a** for a timeline of pathogen circulation based on PCR detections). RSV was the first pathogen to make a strong resurgence and peak in late 2021, followed shortly by heightened seasonal coronavirus activity. A small outbreak of influenza was observed out of season in mid-2022, followed by a larger outbreak during the 2022-23 winter. To estimate the pandemic immunity debt to endemic viruses, we collected convenience serum samples from 999 children <11 yo during three cross-sectional time periods in 2020-2022 and from 509 adults during two paired time periods in 2021-2022 (see **Fig. 1b** for a timeline of serology sampling and methods for details on sampling scheme). The mid-2020 sampling time occurred after a mostly uninterrupted winter season of viral activity in 2019-2020, so that antibody levels in the population would be expected to reflect those of a normal winter season. Subsequent sampling times were expected to capture putative drops in antibody levels as activity of endemic viruses declined. Antibody responses to all seasonal influenza subtypes, RSV, and seasonal and pandemic coronaviruses were measured using the recently developed quantitative Meso Scale Discovery (MSD) electrochemiluminescence multiplex immunoassay (see 20 and methods for details).

**Figure 1.**
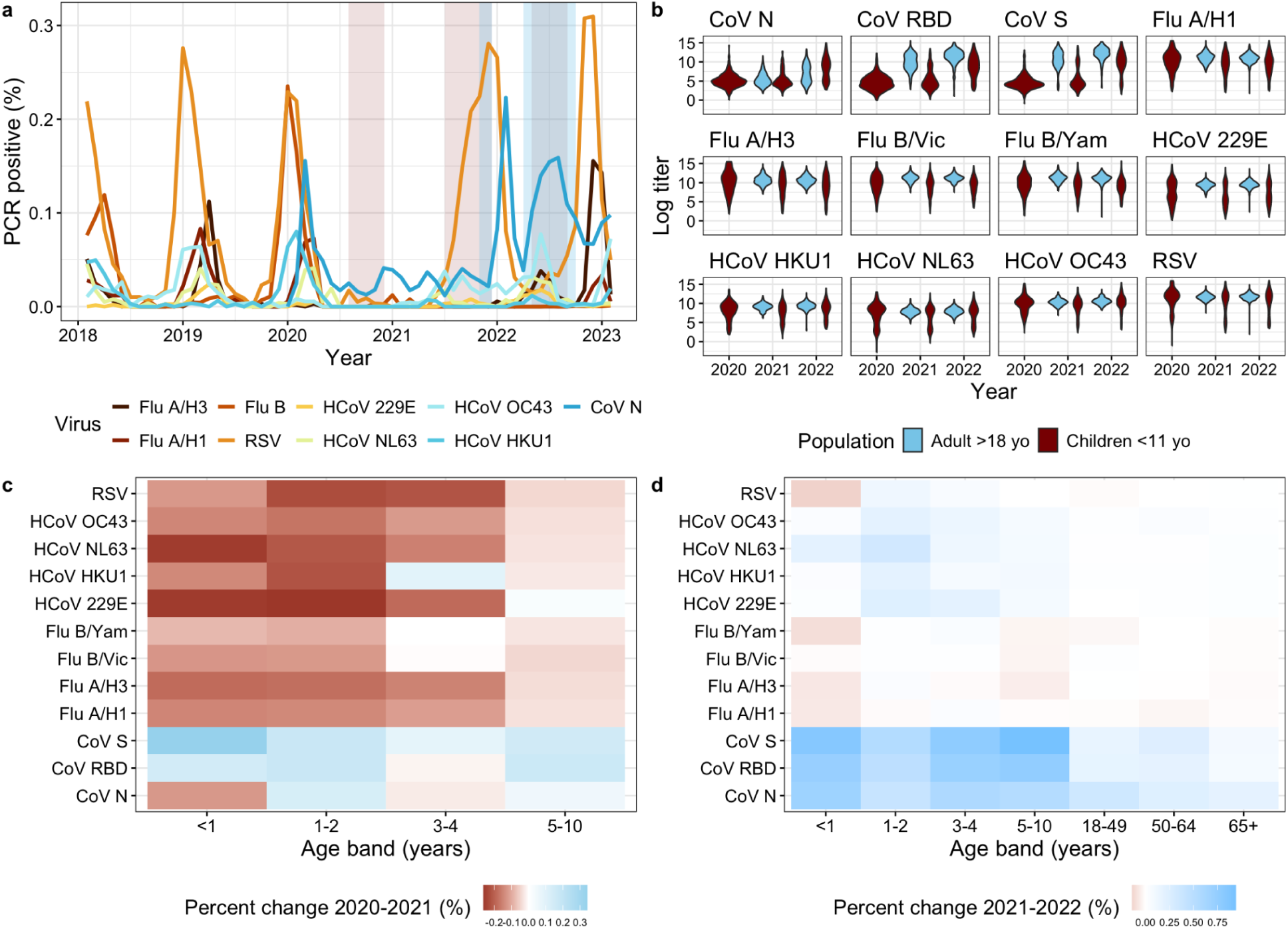
King County respiratory virus surveillance and antibody patterns, 2019-2023. **a)** Time series showing the percent PCR-positive confirmed cases for influenza A/H3, A/H1, B, CoV N, CoV Spike, CoV RBD, RSV, and HCoV 229E, NL63, OC43, and HKU1 reported through Seattle Children’s Hospital and the King County COVID-19 Dashboard. The blue and orange bands represent when serological samples were collected from adults and children respectively. The periods of sampling of children and adults overlap in 2021 and 2022. **b)** Violin plots showing the yearly distribution of raw log_e_ antibody levels for all antigens based on the MSD multiplex assay. Antibody measurements are grouped by adults >18 yo (blue) and children <11 yo (red). **c)** Geometric mean percent change in antibody levels between the 2020 and 2021 sampling points by pathogen subtype for smaller age bands <11 yo, where blue indicates a rise in the mean antibody levels in 2021 compared to 2020 and red indicates a drop. **d)** Geometric mean percent change in antibody levels between the 2021 and 2022 sampling points across the same age and pathogen combinations as in c).

#### Characterizing pandemic immunity debt

We calculated the mean antibody level by age group and pathogen subtype for annual sampling points in 2020-2022. We found that antibody levels declined substantially between 2020 and 2021, particularly for children <5 yo (**Fig. 1c**). The 1-2 yo age group had the most significant declines in antibody levels for all influenza viruses, seasonal coronaviruses, and RSV [**Supplementary I**]. Compared to antibody concentration levels in 2020, children 1-2 yo sampled in 2021 experienced 15.3 - 30.6% relative decreases in antibody levels against seasonal coronaviruses (Kolmogorov-Smirnov p-values < .002), and 17.5% and 18.3% declines against influenza A/H3 (p-value < .005) and RSV (p-values < .0007), respectively. Significant declines in antibody concentration levels were also exhibited for the 3-4 and 5-10 yo age groups for the seasonal alpha coronaviruses and RSV, while <1 yo experienced declines in antibodies to influenza A/H1 and seasonal coronaviruses. In 2022, antibody levels began to steadily increase across most age groups and pathogens, consistent with the resurgence of endemic pathogens observed in surveillance data (**Fig. 1c**). Influenza was the only pathogen for which antibody levels continued to decline in 2022 across several pediatric age groups. (**Fig. 1d**). In contrast to endemic pathogens, antibody concentration levels to different SARS-CoV-2 antigens rose in most age groups throughout 2021-2022, with the largest increase observed among children under 10 yrs following the Omicron wave in 2022.

#### Modeling age-specific immunodynamics

We used Bayesian hierarchical models to infer age-specific antibody kinetics parameters from the King County 2020-2022 serological data (**Fig. 2a-d and methods**) (21). We estimated age-specific differences in the antibody boost and waning rate between children <5 yo and adults for RSV, seasonal coronaviruses, and influenza viruses. We found that children experience larger antibody boosts and quicker waning following pathogen exposure (**Fig 2a-b**). For influenza specifically, we estimated children <5 yo boosted on average 4.5 log arbitrary units per milliliter (AU/mL) (95% CrI: 3.8, 5.7) following A/H3 exposure and waned at a rate of 16% year^−1^ (95% CrI: 5, 27). We estimated that adults boosted on average 1.9 log AU/mL (95% CrI: 1.5, 2.0) after A/H3 exposure, and waned at a rate of 1% year^−1^ on average. These age-specific patterns were also observed for other influenza viruses, RSV, and seasonal coronaviruses. For example, we estimated that children <5 yo experienced a boost of 4.7 log AU/mL (95% CrI: 4.3, 5.1) following infection with RSV, compared to an average boost of 2.7 log AU/mL (95% CrI: 2.5, 2.7) for adults, with children waning annually about 7x more quickly than adults.

**Figure 2.**
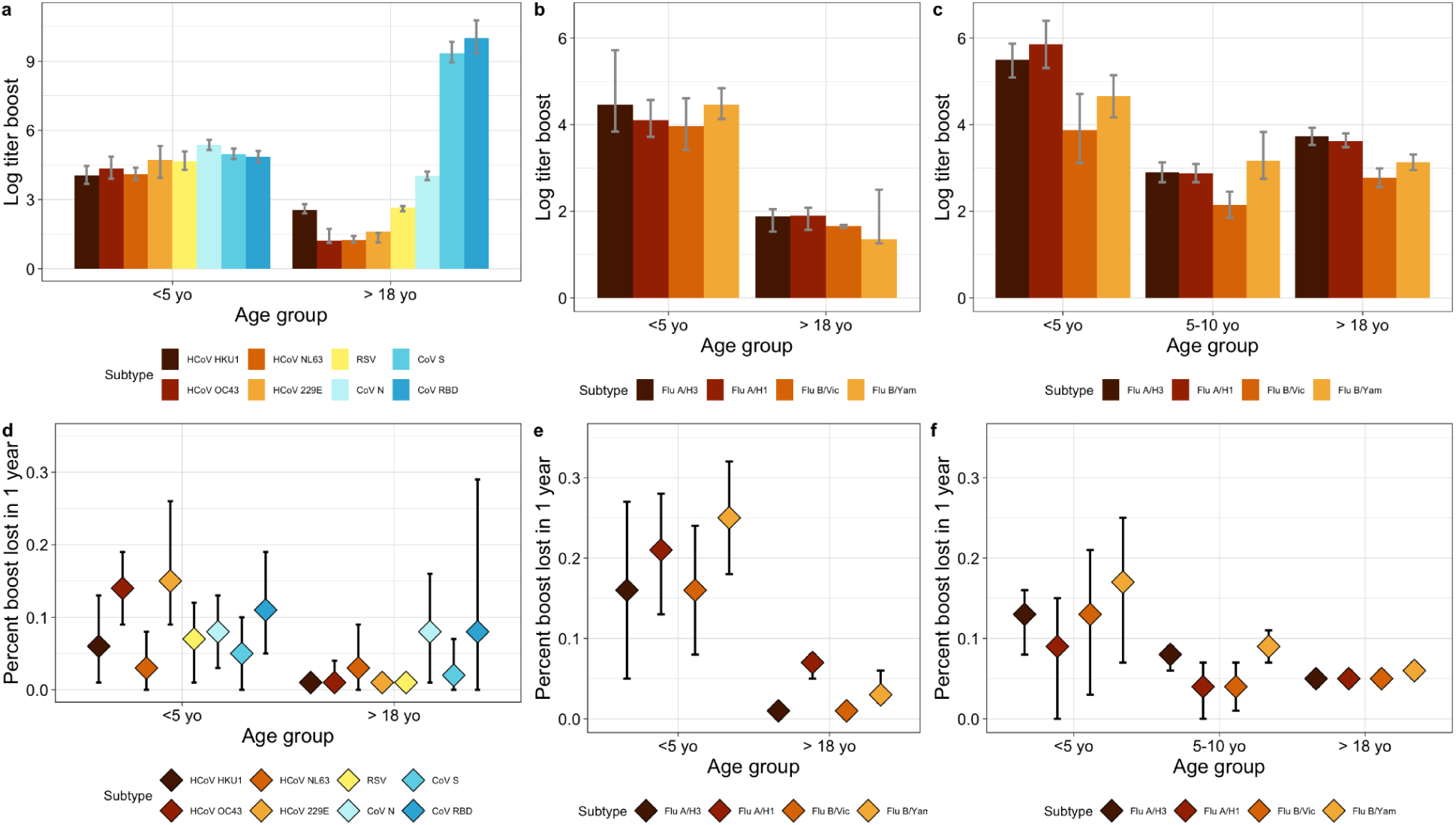
Bayesian hierarchical modeling of age-specific antibody kinetics in King County, 2020-2022, and South Africa, 2016-2018. **Top:** Log antibody concentration boost following infection or vaccination event by pathogen subtype and age group with error bars showing 95% credible intervals. Boosting estimates are presented for **a)** <5 yo and adults in King County for SARS-CoV-2, seasonal coronavirus and RSV, **b)** <5 yo and adults in King County for influenza subtypes, and **c)** <5 yo, 5-10 yo, and adults in South Africa for influenza subtypes. **Bottom:** Estimated percent of the antibody boost lost in one year following infection or vaccination event by subtype and age group with error bars indicating 95% credible intervals. Waning estimates are presented for **d)** <5 yo and adults in King County for SARS-CoV-2, seasonal coronaviruses and RSV, **e)** <5 yo and adults in King County for influenza subtypes, and **f)** <5 yo, 5-10 yo, and adults in South Africa for influenza subtypes. Antibody concentrations and boost estimates are based on the multiplex MSD assay for King County (panels **a) b) d) e)**) and the HAI influenza assay for South Africa (panels **c) and f)**).

To contrast the immunological patterns of endemic viruses with those of a pandemic pathogen to which pre-existing immunity was low or inexistent, we repeated this analysis with SARS-CoV-2 antibodies. We found that adults experienced the highest rates of waning to SARS-CoV-2 nucleocapsid and receptor binding domain (RBD) antigens, waning at a rate of 8% year^−1^ on average (**Fig. 2a**). We also estimated that adults experienced a larger boost to SARS-CoV-2 antigens upon exposure than to any other endemic respiratory virus. Estimated antibody responses to SARS-CoV spike and RBD, which capture composite exposures to vaccination and natural infection, were significantly higher than that of infection-based SARS-CoV-2 nucleocapsid, with estimated boosts of 9.3 (95% CrI: 8.9, 9.8) and 10.0 (95% CrI: 9.3, 10.8) for the spike and RBD antigens respectively, compared to 4.0 (95% CrI: 3.8, 4.2) for nucleocapsid. Waning of SARS-CoV-2 nucleocapsid antibodies did not differ between children and adults.

#### Immunodynamics of influenza in an independent population

To further investigate the age differences in immunological dynamics evidenced in the King County data and confirm our findings using a different serological assay, we turned to a companion study from South Africa designed to monitor influenza immunity in a high-transmission setting (22). In this household cohort study, paired samples were available for 375 children and 653 adults who were monitored for at least a year during 2016-2018 (**Fig. 2c, 2f**). Samples were tested by the hemagglutinin inhibition assay for all influenza subtypes and using strains circulating during the study period. Modeling antibody levels in this cohort revealed similar age immunodynamics than in the King County study, with children <5 yo of age experiencing significantly higher antibody boosts following infection and quicker rates of waning. Across all four influenza subtypes, we estimated that on average children waned at a rate of 9-17% year^−1^, compared to 5-6% year^−1^ for adults. We also found that children 5-10 yo mirrored the immunological dynamics exhibited by adults more closely than those of children <5 yo, suggesting a fundamental change in immunological response or risk of exposure beginning around the age of five. Age-specific influenza patterns were similar between the King County and South Africa populations despite vastly different vaccination rates: 39% of children were vaccinated against influenza in King County, whereas none were vaccinated in South Africa.

### Population-level influenza transmission model informed by serology

Next, we linked our serological insights obtained from individual-level data to realized epidemiological patterns on a population level. We tested whether the age differences in immunodynamics evidenced in our serological data could reproduce key features of the post-pandemic pathogen rebound in Seattle. We calibrated a mechanistic disease model to monthly age-structured time series of influenza hospitalization and emergency department visit data (referred to here and after as healthcare encounter data) from King County, WA, from 2017-2023 (**Fig. 3a**). Healthcare encounter data serves as a useful marker of age-specific changes in disease patterns throughout the pandemic. Compared to three pre-pandemic seasons, the 2022-23 influenza rebound season exhibited slightly earlier timing and larger magnitude. There were also age structure shifts, where individuals 5-19 yo comprised 32.5% of the healthcare encounter burden in the 2022-23 rebound season, compared to 24.4% across the pre-pandemic seasons (Chi-square p-value < 2.25e-74) (**Fig. 3b**).

**Figure 3.**
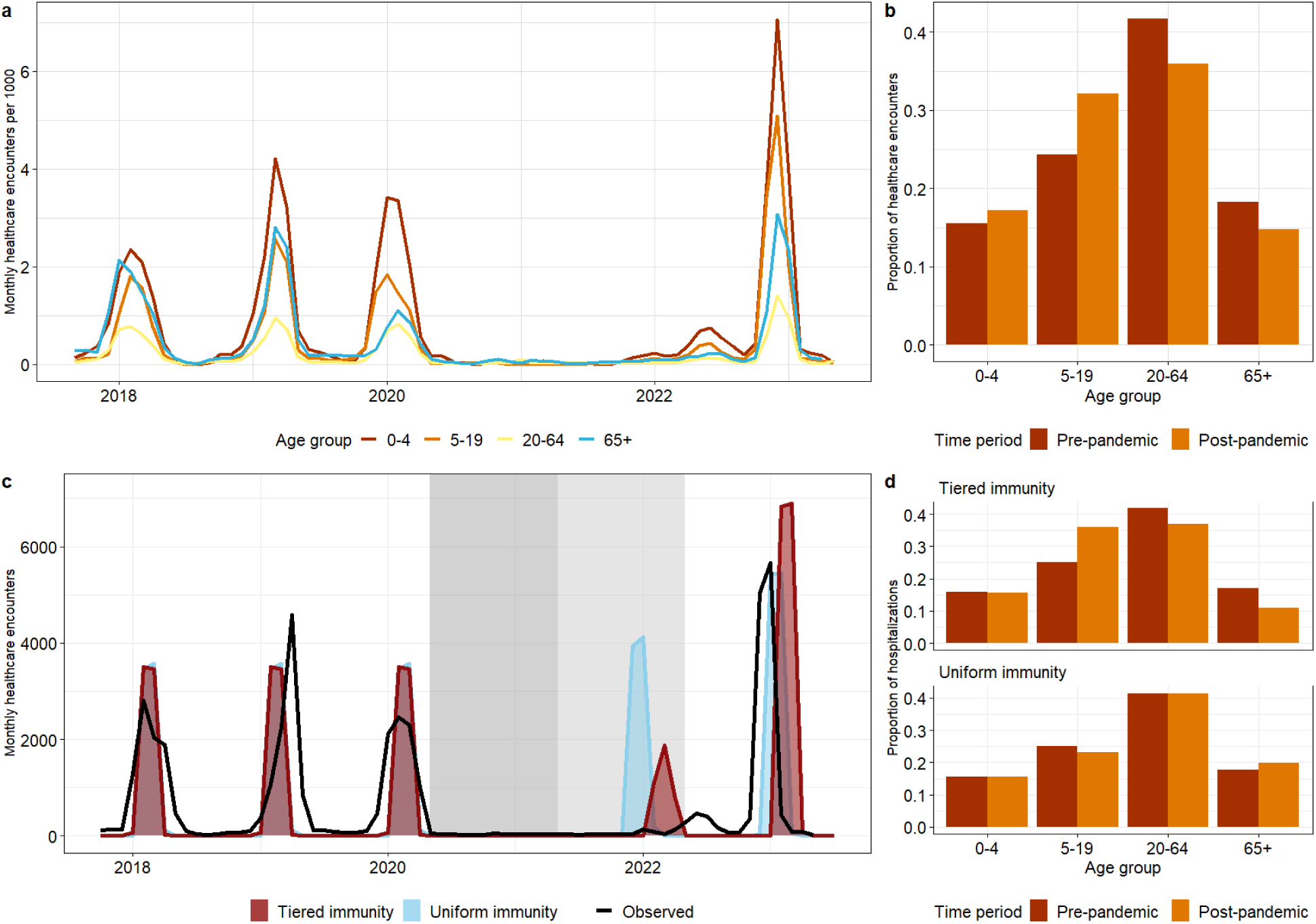
Age-specific influenza healthcare encounter time series and predictions by transmission model, King County, WA, 2017-2023. **a)** Monthly aggregated healthcare encounters per 1000 persons segmented by age into the following groups: 0-4 yo, 5-19 yo, 20-64 yo, and 65+ yo. **b)** Observed shift in influenza healthcare encounter age structure, where pre-pandemic represents the 2017-2020 seasons and post-pandemic represents the 2022-23 season. Bars show the proportion of healthcare encounters in each age group out of reported healthcare encounters. **c)** Predicted age-structured healthcare encounters for a model assuming tiered immunity (where loss of immunity is allowed to differ by age, blue) and a model assuming uniform immunity (immunity does not vary by age, orange). Each model was fitted to the observed incidence time series (represented by the solid black line for the entire population). Fitting of the tiered immunity model suggests that children under 5 yo lose immunity at a rate twice faster than older individuals. **d)** Comparison of the predicted age structure of the tiered immunity model (top) and the uniform immunity model (bottom) for the pre- and post-pandemic periods (red and orange, respectively).

To evaluate the value of our serological insights to explain population-level transmission dynamics, we designed two age-structured compartmental models with different immunity structures (see methods for details). Inspired by the waning patterns in the King County and South African serology data, the first model assumed a “tiered immunity” structure stratified by age, where the relative difference in the rate of immune waning between individuals under and over 5 yo was a free parameter. We then considered a second “null” model where we assumed the same duration of influenza immunity across all age groups (“uniform immunity” model). We fitted each of these models to the King County monthly age-specific healthcare encounter time series from 2017-2023 to test the impact of incorporating age-specific immunity. We allowed for two periods of reduced transmission in line with NPIs implementation from March 2020-March 2021 and April 2021-April 2022. Parameters such as the influenza transmission rate, seasonal forcing, relative difference in rate of immune waning by age (for the tiered immunity model), the effect of social distancing on transmission, and the proportion of infections resulting in healthcare encounters were estimated, while other natural history parameters were drawn from the literature (see Appendix Table 4 for model parameters).

Based on calibration of the tiered immunity model to epidemiological data, we estimate that children <5 yo experience waning at about twice the rate of individuals over 5 yo, losing full immunity after about 2 years (Appendix Table 4). Calibration further revealed differences in the estimated effects of NPI between the tiered and uniform immunity models. In the homogeneous immunity model, we estimated that NPI reduced transmission by 43% and 27% for the first and second NPI periods. In the tiered immunity model, these parameters were 50% and 44% respectively. We assessed the root mean square error for each model and found similar performance between model structures (methods). However, models differed in the predicted dynamics of the influenza rebound. The tiered immunity model more accurately reproduced the timing of the first out-of-season outbreak, particularly capturing the observed lack of circulation in 2021 and early 2022 (**Fig. 3c**). Both models predicted a later rebound in 2022-2023 compared to observations, although the uniform immunity model was slightly more accurate with respect to timing. The overall magnitude of the 2022-23 rebound season was similar by the two models, with the tiered and uniform immunity models predicting overall healthcare encounter burden to be 115% and 91% of what was observed. Differences in the predicted age structure of healthcare encounters were more marked. While both models predicted the pre-pandemic age structure appropriately, predictions for the post-pandemic rebound differed. Only the tiered immunity model accurately predicted the post-pandemic age shift observed in the data, where the proportion of encounters increased from 22 to 33% between periods in the 5-19 yo, with a proportional decrease in older age groups (**Fig. 3d**). The observed data showed a slight increase in the proportion of healthcare encounters in <5 yo from the pre- to post-pandemic, which was not captured by either of the models.

## Discussion

The COVID-19 pandemic period provides a unique opportunity to observe antibody dynamics in the absence of circulation of endemic pathogens. We analyzed serological data generated from a novel multiplex assay in the first three years of the pandemic in King County WA, USA. We found a substantial antibody debt among children but not in adults in 2021 across a diverse array of pathogens, including seasonal coronaviruses, influenza viruses, and RSV. Further, the antibody debt disappeared among children as pathogen circulation rebounded in 2022. Modeling of antibody kinetics revealed greater antibody boosting and quicker waning in children under 5 yo compared to adults, and this pattern was consistent across endemic viruses. Using auxiliary influenza serology from South Africa, we demonstrated that these immunological patterns were upheld in a contrasting, non-pandemic and unvaccinated setting, and using a different serologic assay. Integrating these age immunodynamics in a population-level model helped capture the observed epidemiology of the influenza rebound in King County, particularly a shift in the age distribution of healthcare encounters towards older children. Our serological findings, in combination with population-level modeling, support that age-specific waning, particularly among children, may be an overlooked but important driver in the outbreak dynamics of influenza and other seasonal respiratory viruses.

A prior influenza serological analysis set in the pre-pandemic period found differences in antibody kinetics by age, showing that children <15 yo waned at a modestly quicker rate to influenza A/H3 than adults (17). Our analyses extend these findings to other pathogens and suggest that antibody waning may be further pronounced in younger children <5 yo. A recent serologic study from Norway noted a marked drop in antibodies to influenza A/H3N2 and A/H1N1 in children under 5 yrs by summer 2022, while antibody levels remained more stable in older age groups (23). Another serology study in the Netherlands supports a slow rate of antibody waning to influenza during the COVID-19 pandemic among adults, with little evidence of an immunity debt in this population (24), aligning with our serological findings in adults. Taken together, these results support a post-COVID19 pandemic immunity debt primarily concentrated among children. More broadly, the observed age differences in immunodynamics lend support for collection and analysis of serologic information across a range of age groups, and particularly among young children.

Our findings are consistent with what is known about the role of age in antibody dynamics. Upon first exposure to viral infections in childhood, the immune system mounts a large and broad antibody response, from which differentiated memory B cells are subsequently generated (25,26). Later in life, exposures elicit a more specific antibody response, often associated with smaller rises in antibody concentration levels upon infection (15,27). These patterns are a feature of age but also exposure history, as evidenced by SARS-CoV-2 data in our study. In King County adults, antibody responses to the nucleocapsid antigen (a marker of natural infection) were characterized by a relatively larger boost and quicker waning than to other pathogens, mirroring the antibody kinetics typically exhibited by children <5 yo to endemic pathogens. Larger boosts, commonly associated with lower starting antibody concentration levels, are not necessarily correlated with increased protective immunity but may instead be a marker of a novel exposure (21).

Many studies have established that children are re-infected with influenza and other respiratory viruses more frequently than adults (28–31). There are several potential explanations for this, including few prior pathogen exposures, higher contact rates, and less mature respiratory and cardiovascular systems (32,33). Our analysis suggests that quicker waning of immunity, specifically immunoglobulin G (IgG) antibodies, may contribute to shaping these age-specific dynamics. For influenza, there are known to be age-specific differences in epitope targeting that may partially explain these patterns, whereby a child’s antibody response is directed towards the HA head, while adults target the more conserved stalk and other epitopes, which are associated with longer-lasting, protective immunity (34). This may only partially explain our findings given that age-specific differences in waning were observed for pathogens with lower rates of antigenic evolution than influenza. Regardless, prior analyses found that antibody concentration levels were predictive of a child’s susceptibility to infection (17), suggesting that the rapid loss in antibody concentration levels that we measured in young children may result in loss of protective immunity. However, we acknowledge that in the absence of PCR-confirmed infection data from the same individuals that were serologically sampled in King County, we cannot make definitive assertions regarding how pre-existing antibody concentration levels relate to protective immunity.

To link our serological insights to realized population-level epidemic patterns, we fitted a transmission model to healthcare encounters reported during the pre- and post-COVID-19 period and assumed that a reduction in antibody concentration levels results in a loss of protective immunity. We found that models assuming different age-specific immunity structures differed in their predictions of the timing and age structure of healthcare encounters in the large influenza rebound season in 2022-23. When we considered age-specific differences in waning of protective immunity, we were able to accurately capture the reported shift in healthcare encounters towards children 5-19 yo during the rebound season. This age shift was likely driven by a cohort of young children who aged out of the <5 yo age class during the COVID-19 pandemic without gaining primary or secondary exposure to influenza. Combined with a fast waning in this age class, these children would have in turn fueled a pool of susceptible hosts in the 2022-23 rebound season as they aged into the 5-19 yo age group. In contrast, a transmission model assuming a fixed pace of immune waning in all age groups cannot fully explain the timing and age dynamics of the influenza rebound season. It is also worth noting that modeling of serologic data and population-level incidences independently support a rate of immune waning twice faster in young children compared to adults. Overall, our results indicate that consideration of age-specific immunodynamics can improve the prediction accuracy of models of respiratory virus outbreaks.

Our transmission modeling analysis revealed that accounting for age-specific immunity resulted in a lower estimated seasonal forcing parameter, in turn necessitating a greater impact of NPIs to limit transmission during the COVID-19 pandemic period. Prior modeling analyses have shown that environmental factors likely mediate influenza transmission dynamics but cannot fully explain the variability in the timing of seasonal outbreaks (35,36), with inter-annual fluctuation in prior immunity and seeding accounting for some of this variation. This may have important implications for the control of seasonal influenza and potentially other endemic respiratory pathogens as well. Given the age-specific dynamics observed, better serologic monitoring of immunity trends in children may be useful for predicting outbreaks and intervention planning (37). The high numbers of contacts experienced by children relative to other populations (33) accentuates the importance of monitoring child immunity to anticipate population-level dynamics.

Our findings of differential antibody dynamics in young children were consistent in two different populations in the US and South Africa and supported by different serological assay measurements. In South Africa, we studied paired sera originating from an unvaccinated population throughout several pre-pandemic seasons (22). In King County, WA, we had access to cross-sectional data for children <11 yo, and our population was highly vaccinated. Exploratory analysis showed slightly elevated titers to influenza antigens for children from King County who had recently received an influenza vaccine, compared to unvaccinated children (not shown). However, the drop in influenza antibody titers between 2020 and 2021 was similar (17-29% for vaccinated vs 15-23% for unvaccinated). While we can’t fully disentangle the role of natural immunity and vaccination in our King County analyses, we note that age differences in the pace of antibody waning were maintained in an unvaccinated population in South Africa. Another noteworthy difference between the two populations was the higher estimated influenza attack rates in South Africa, likely driven by population demographics [**Supplementary II**]. Overall, the age-specific immunodynamics reported here may reflect a universal phenomenon that is robust to extrinsic factors that affect the intensity of influenza circulation, such as pandemic perturbations, population structure and contacts, and vaccine interventions.

Several critical distinctions between the assays used to analyze the US and South African influenza serology are worth noting. First, the MSD and HAI assays measure different biomarker responses, so that output measurements cannot be compared directly. The HAI assay used to analyze the South African serology measures the extent to which antibodies inhibit the binding of the HA surface protein to red blood cells, which has been shown to play a critical role in virus neutralization across age groups. In contrast, the MSD assay measures a broader antibody-binding response targeted to the HA protein which is less specific to a particular influenza strain (28,38). This means that the HAI assay is more likely to detect responses specific to the antigenic component of an influenza strain, while the MSD assay is more likely to capture cross-reactive immune responses to a range of prior circulating strains of a given subtype. Further, the MSD assay is more sensitive than standard assays, thus likely capturing more transient boosts following exposure (39). The consistency of our MSD findings across several respiratory virus pathogens for which no cross-reactivity would be expected, combined with the specificity of the HAI influenza assay, suggest that the observed age-specific immunodynamics are common to a range of subtype- and strain-specific respiratory virus antibodies.

There are several methodological limitations worth considering. We did not account for potential cross reactivity between strains of a given pathogen. Given the broad response captured by the MSD assay, we expect to observe cross reactivity within subtypes of the studied pathogens. We have assessed within-individual correlation by age and note this will be an important area of future work [**Supplementary III**]. The fact that our findings are corroborated with the more specific HAI assay for influenza suggests that cross-reactivity does not play a major role in our estimates. Second, in the Bayesian modeling framework, we informed the prior on attack rates for all respiratory pathogens based on the distribution of annual influenza attack rates observed in the South African cohort study, since we did not have representative incidence data from King County, WA.

Our transmission modeling analysis also has several limitations. The number of reporting healthcare facilities increased over time in King County. Because we did not have information on how the overall catchment size increased throughout the study period, we scaled the healthcare encounter time series linearly in accordance with the number of reporting facilities. Further, there could have been differences in testing or reporting rates by age group, which may have contributed to the observed age shift. Although this is an imperfect adjustment to the time series, we compared our healthcare encounter time series with the US CDC’s influenza-like illness Washington-specific dataset to ensure that the overall nature of the pre-pandemic and post-pandemic rebound are captured [**Supplementary IV**]. Further, we used an age-structured SEIRS influenza model which combines all influenza subtypes. The rebound season in 2022-23 in the US was marked by predominance of influenza A/H3N2 subtype (70%), with co-circulation of A/H1N1 (30%) (40). Relatedly, we did not consider the role of vaccination or antigenic evolution. While antigenic novelty can affect inter-annual fluctuations in influenza incidence (41), the impact on the age distribution of cases remains debated (42). Moreover, while the tiered immunity model had a better fit to the age distribution of cases in the large rebound season, neither the tiered or uniform immunity models fully reproduced the reported epidemiological dynamics. Because we aimed to make inferences about broad immunodynamics conserved across diverse locations, where vaccination and strain cycling may differ, and our epidemiological time series data was limited, we chose a simple model structure to investigate our primary question qualitatively. Lastly, inspired by antibody patterns in the serology data, we tested one of several hypotheses for the shift in influenza dynamics post COVID-19, namely the role of age-specific waning in protection against infection. Influenza immunity is complex and we cannot rule out the contribution of other immune responses that are not modulated by antibodies and confer protection against severe disease. Future work should investigate the impact of parameterizing age-specific duration of immunity against infection and severe disease in more complex influenza models, especially as waning dynamics may depend on the type of immune functions, influenza subtypes, vaccination, and antigenic changes.

We have confirmed that immunity debts were observed throughout the COVID-19 pandemic in children, as evidenced by drops in antibody concentrations across several respiratory pathogens in the early pandemic period. Antibody kinetics measured from two different assays support substantial differences in immune response to respiratory pathogens in children under the age of five compared to older individuals. A transmission modeling analysis indicates that age-specific immunodynamics may impact overall circulation patterns, particularly amidst pandemic perturbations. Future work should focus on elucidating the mechanisms responsible for the observed age-specific immunodynamics and determine how increased serological surveillance efforts could improve targeted public health measures.

## Methods

### Data

#### I. Serology and viral activity in King County, WA

Our serology study was initiated in November 2021 as an add-on to the larger Seattle Flu Alliance surveillance study (SFA), which began in 2018 and carried out hospital and community-based surveillance to monitor the circulation of 26 respiratory pathogens (43). The purpose of the serology arm of SFA was to document the immunity debt that may have accumulated in the first two years of the COVID-19 pandemic and explore changes in antibody levels coinciding with putative pathogen rebound in the 2021-22 winter. Cross-sectional sera were collected from convenience residual blood samples of 999 immunocompetent children seeking medical care, evenly distributed between the <5 yo and 6-10 yo age groups, through Seattle Children’s Hospital. We also sourced convenience residual sera from a blood donor bank in Seattle, Bloodworks Northwest, which collected specimens from 509 adults prospectively, 274 of which provided repeat samples (44). We collected the same number of samples in two adult age groups, 20-64 and over 65 yo. We defined three sampling periods to titrate immunity at key points of the pandemic, after balancing the availability of samples and seasonality of pathogen circulation (**Table 1**). The first period, occurring in the summer of 2020, followed a mostly regular respiratory season in 2019-2020, with only samples from children <11 yo being collected. In subsequent periods, sera was collected from both adults and children. The second period in fall 2021 followed a year and a half of repressed pathogen circulation due to COVID-19 interventions and was presumed to mark a trough in immunity. The third period in spring-summer 2022 followed a respiratory virus season in which multiple pathogens rebounded (**Figure 1A**).

**Table 1:**
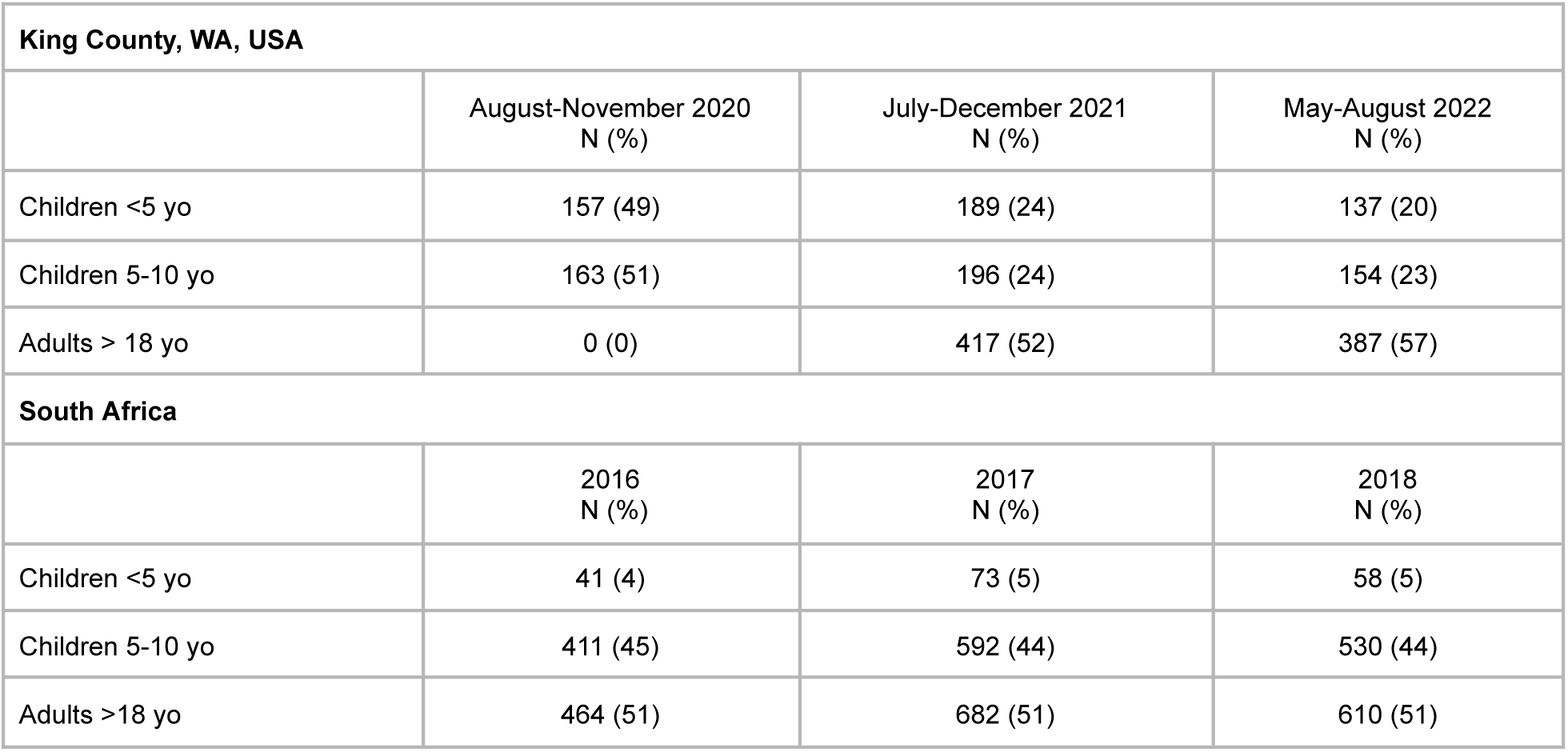
Serological sampling by location, age group, and time period. The number of unique samples analyzed annually for King County, WA, USA 2020-2022 and South Africa 2016-2018. Both populations are disaggregated by age groups <5 yo, 5-10 yo, and adults. More detailed tables showing sample sizes by pathogen are provided in **Supplementary II**.

Sera was analyzed using the multiplex Meso Scale Discovery (MSD) electrochemiluminescence immunoassay and laboratory analysis was performed at the Fred Hutchinson Cancer Center (20). We used the V-PLEX COVID-19 Respiratory Panel 3 IgG Kit to analyze antibody concentration in arbitrary units per milliliter against Flu A/Hong Kong/2014 H3, Flu A/Michigan/2015 H1, Flu A/Shanghai/2013 H7, Flu B/Brisbane/2008 HA, Flu B/Phuket/2013 HA, HCoV-229E Spike, HCoV-HKU1 Spike, HCoV-NL63 Spike, HCoV-OC43 Spike, MERS-CoV Spike, RSV Pre-Fusion F, SARS-CoV-1 Spike, SARS-CoV-2 N, SARS-CoV-2 S1 RBD, and SARS-CoV-2 Spike antigens (SARS-CoV-2 based on ancestral strain). We transformed antibody concentration to log_e_ units. The sample collection and this study were approved by the Institutional Review Board of Seattle Children’s Hospital. This study was granted a waiver of consent since it used residual clinical samples and existing clinical data.

To provide context for serology, we sourced detailed respiratory surveillance data from Seattle Children’s Hospital (SCH) from January 2018-January 2023. SCH performed 105,207 PCR tests among children <21 yo in the inpatient and outpatient setting during this time period. Children were tested for influenza and RSV using the Cepheid Xpert XpressR (Sunnyvale, CA) panel and for influenza A/H3N2, A/H1N1, HCoV 229E, NL63, OC43, and HKU1 using the Biofire FilmArray^R^ multi-pathogen respiratory viral panel (Biomerieux, Salt Lake City, UT). We present monthly tests percent positive for these pathogens to visualize endemic respiratory pathogen circulation throughout the study period and additionally derive a COVID-19 time series from the King County COVID-19 Dashboard (45). Because we were interested in how serology could help understand the influenza rebound in the post-COVID-19 pandemic period, we obtained more resolved age-specific data on influenza healthcare encounters from Public Health - Seattle & King County (PHSKC). The dataset comprises age-specific (0-4, 5-19, 20-64, over 65 yrs) emergency department visits and hospitalization data reported by PHSKC to the Washington State Department of Health Rapid Health Information Network (RHINO) (46). The number of King County hospitals that report to RHINO increased from 6 to 21 from 2017-2022. Given limited information on how the underlying surveillance catchment area changed, and to obtain a stable time series of influenza healthcare encounters, we assumed that reporting increased linearly with the number of reporting hospitals. This means that the time series was upscaled by the fraction of reporting hospitals relative to the final set of 21.

#### II. Serology in South Africa

Because we used a novel multiplex serological assay to assess immune dynamics in the King County, WA, population, we looked for confirmation of our findings with an independent dataset and assay. Accordingly, we sourced serologic data from a 3-year household cohort of influenza in South Africa (22,47). We analyzed paired sera collected from 1684 individuals through the *Prospective Household cohort study of Influenza, Respiratory syncytial virus, and other respiratory pathogens community burden and Transmission dynamics* (PHIRST) in a rural and urban setting located in Mpumalanga and North West Province, South Africa, 2016-2018. Detailed study protocols have been published elsewhere (22,47). Longitudinal samples were collected at study enrollment and before and after the influenza season annually in 2016, 2017, and 2018 for enrolled households, including 375 children and 653 adults (**Table 1**). Antibody levels were measured using the well-established hemagglutinin inhibition assays against influenza strains circulating during the study period, namely A/South Africa/2517/2016 EPI_ISL_230453, A/Singapore/INFIMH-16-0019/2016 EPI_ISL_225834, B/South Africa/R3037/2016 EPI_ISL_231726, and B/South Africa/R5631/17 EPI_ISL_17008503. Antibody concentration was transformed into log_2_(x/5) units.

##### Serological analyses

We first used simple descriptive statistics to assess changes in antibody concentration levels for individuals in King County by age group and pathogen throughout the study period. We summarized the serological data by 7 age groups: <1 yo, 1-2 yo, 3-4 yo, 5-10 yo, 18-49 yo, 50-64 yo, and 65+ yo, and calculated the percent change in average antibody levels between the 2020, 2021, and 2022 time points. Two-sample Kolmogorov-Smirnov tests were performed to assess whether the distribution of antibody measurements by age band and pathogen changed across annual time points [**Supplementary I**].

Next, we used a Bayesian hierarchical model developed to make epidemiological inferences from individual-level serological data using the R package *serosolver* (21). The model estimates the joint posterior distribution of (1) underlying antibody kinetics following infection, including antibody boosting and waning, (2) infection histories for all individuals throughout the study period, and (3) population infection probability over time. In short, the model estimates the set and timing of infections that are most consistent with an individual’s antibody profile, accounting for dynamic antibody responses following infection and measurement variability. We separately fit the model to the King County serological data using cross-sectional data from children <5 years and paired sera from adults for RSV, SARS-CoV-2 N, the seasonal coronaviruses HCoV 229E, NL63, OC43, and HKU1, and influenza subtypes A/H3N2, A/H1N1, B/Victoria, and B/Yamagata (**Table 1**). We did not model the 5-10 yo age group cross-sectional data due to the challenge of distinguishing between high initial antibodies and antibody boosts following exposure (28). We also excluded infants <6 months old to ensure that our results were not biased by transplacental maternal antibody dynamics in young infants (48). [**Supplementary V**].

We additionally fit the same model to paired serological samples from South Africa for three age groups, <5 yo, 5-10 yo, and 18+ yo, and for each influenza subtype: A/H3, A/H1, B/Victoria, and B/Yamagata (**Table 1**). The model was fit to two samples per individual in the South African population to maximize sample size.

We adopted a simplified version of the *serosolver* modeling framework, where the observed antibody level *Y* for individual *i* at a given time *t* for age group *a* is the sum of antibody boosting µ_s_ that has accumulated since birth and wanes at rate *r* following exposure. We assume that estimated parameters are pathogen- and subtype-specific (*p*). An individual’s expected antibody level can then be modeled as:

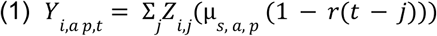

where the distribution of observed antibody levels for a given age group and pathogen is normally distributed (truncated give the upper and lower limit of detection) with mean *Y_i, a, p, t_* and the set of antibody kinetics parameters to be estimated is defined by Θ = { µ_*s*_, *r*, ***σ*** }, where ***σ*** is the estimated measurement error. Individual infection histories are represented as vectors of binary latent states *Z*_*i,j*_ indicating whether an individual was infected (*Z*_*i,j*_ = 1) or not infected (*Z*_*i,j*_ = 0) in a given time period. We estimated infection histories back to birth with the exception of SARS-CoV-2 antigens, which are estimated from 2020-2022. The model is run at an annual timescale, meaning an individual can be infected a maximum of once per season. Each infection event *Z*_*i,j*_ is modeled by a Bernoulli trial that is conditional on the time-varying population probability of infection ϕ.

The *serosolver* framework allows for the implementation of different priors on the probability of infection ϕ depending on the specific disease context. We implemented a beta prior on the probability of infection in each period *t*, which is appropriate when an individual’s time-varying probability of infection is governed by the force of infection; as is the case for endemic respiratory pathogens (21). In this model framework, the per-capita attack rate is beta-distributed and the total number of infections experienced by each individual follows a binomial distribution. To set the priors, we inferred the distribution of influenza population attack rates from PCR-confirmed data collected through the PHIRST study. Annual attack rates were right-skewed with a mean of 10% standard deviation of 13%; we used this to define our infection probability prior for all endemic respiratory pathogens across both study locations. Thus the population probability of infection *P(*ϕ) is defined by a beta distribution in which we have set alpha = 0.37 and beta = 3.5 (mean: .10, standard deviation: .13) [**Supplementary V**].

The joint estimation of all parameters given observed individual-level antibody data is defined in equation 2:

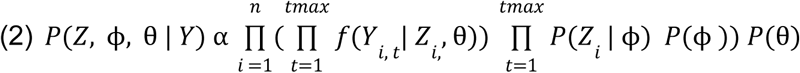

The model uses an adaptive Markov Chain Monte Carlo (MCMC) Metropolis-Hasting algorithm to infer *P*(*Z*, ϕ, θ | *Y*). We performed each model run with a minimum of four chains and 500,000 iterations per chain and assessed post-run model convergence by ensuring *R̂* was < 1.1 for all parameters in each model run.

#### Population-level influenza transmission model informed by serology

Next, we integrated our serologic findings into a mechanistic transmission model to assess how age-specific immunodynamics may impact transmission, particularly in the post-COVID-19 period when endemic pathogens rebounded. We focused this analysis on influenza for which the age dynamics of the rebound in 2021-23 was particularly poorly explained (19) and for which we had more serological data. We considered two variants of a Susceptible-Exposed-Infected-Recovered-Susceptible (SEIRS) model, a null model in which we assumed uniform duration of immunity across the population, and a tiered immunity model, in which we assumed that children <5 yo experienced a different waning rate than older individuals (to be estimated), inspired by our serological analysis. We calibrated these models to monthly age-specific influenza healthcare encounter data for King County, as reported to the RHINO’s syndromic surveillance system which collates visits based on discharge diagnosis codes. The full set of model equations is given by equations 3-6:

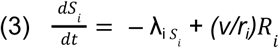

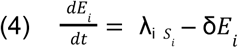

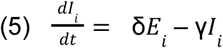

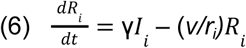

where *S* is the susceptible population; *E* is exposed; *I* is infectious; *R* is recovered, and *i* represents a given age cohort (monthly age cohorts for children ≤ 5 yo and 5-year age cohorts for persons aged 5-75 yo). 1/γ is the infectious period, which is set to 2.27 days, and 1/δ is the latent period, which is set to 2 days based on literature (49–53).

To integrate age-specific immunodynamics in this model, we explored different age-specific immunity durations. Let 1/𝑣 be the baseline duration of immunity and r_i_ the proportion change in duration of immunity for age *i* compared to baseline. We fix 1/𝑣 = 4 years based on prior literature (17,54,55). In the tiered immunity model, we allow the proportion change in immunity duration *r_i_* to vary in children under 5 yrs, while retaining the longer immunity duration in older individuals (ie r_i_=1 for all ages above 5 yrs, so that duration of immunity is 4 years for individuals > 5yo). We then consider a range of r_i_ values that capture putative durations of immunity in children < 5 yo that are both longer and shorter than in older individuals and estimate the value of r_i_ that is most consistent with the incidence data at the calibration stage. In the uniform immunity model, r_i_ is set at 1 no matter the age group (i.e., immunity lasts on average 4 years for all ages).

*λ_i_* i the influenza force of infection on a susceptible individual in age class *i and* is given by equation 7:

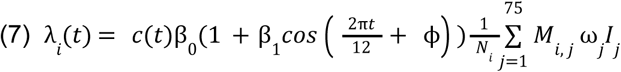

Here *t* is time in months, λ_i_*(t)* is the monthly transmission rate, β_0_ is the baseline transmission coefficient, and *β*_1_ and ϕ represent the amplitude and phase shift respectively that capture the seasonality in transmission in King County, to be estimated from the data. *M_i,j_* describes the contact matrix between individuals in age groups *j* and *i.* We used an expanded version of the contact matrix originally described by Mossong et al., which describes population mixing patterns for several European countries (56). ω*_i_* represents different infectiousness by age, with children under 5 years assumed to be 2x more infectious than older age groups (57). *c(t) is* the time-varying strength of the pandemic-related control periods, representing the percent reduction in transmission due to NPI, where 0 < *c(t)* < 1 and c(t) is to be estimated at calibration stage.

Our model generates the number of new infections, which is given by δ*E_i_*, while our observations are healthcare encounters. To connect the two, we write that Hi∼ Poisson(h_i_δ*E_i_*), where Hi is the number of reported healthcare encounters at time t in age group i, δ*E_i_*, is the number of new infections at time t, and h_i_ is the proportion of infections in this age group that result in reported hospitalizations, where h_i_ is to be estimated from the data. The full parameter table can be found in **Supplementary VI**.

We calibrate the uniform and tiered immunity models in two steps. In the first step, we calibrate the model to age-specific influenza infections for the pre-pandemic period (October 2017-March 2020). In the absence of observed population-level influenza infection time series, we create synthetic time series based on reported healthcare encounters. We estimate that in a given influenza season, 25-35% of children and 15% of adults are infected (29,58–60). Using the population size of King County in 2021, we scale up the observed age-specific monthly healthcare encounter time series to align with realistic incident infection curves, creating a synthetic infection time series that reflects both the true seasonality trends captured by the healthcare encounter time series and realistic population attack rates. To calibrate the baseline uniform immunity model, we fit the transmission parameters (*β*_o,_ *β*_1,_ and ϕ) and age-specific hospitalization rates h_i_. In the age-specific tiered immunity model, we fit one more parameter, the relative duration of immunity in children vs adults *r_i_*, to test whether there is support in the data for a different (shorter or longer) duration of immunity in children.

In the second calibration step, we fit two control periods beginning in March 2020 to estimate the effect of NPIs on influenza transmission in the first two years of the pandemic. We are particularly interested in how these NPI affect the influenza rebound in the 2022-2023 season, where influenza hospitalization burden was higher than expected, particularly in children. We allowed for two different NPI periods to account for differences in overall transmission impacts of the first year of the pandemic (March 2020-March 2021), when more stringent NPIs were implemented in King County, and the second year (April 2021-April 2022), ending when Omicron-related precautions were subsiding. All parameters were estimated using maximum likelihood. We also calculated the root mean square error (RMSE) on age-specific hospitalization time series to assess the fit of the uniform and tiered immunity models (RMSE=3792 vs 3920 respectively). Lastly, we performed a Chi-square test to analyze changes in the age distribution of observed influenza healthcare encounters between the pre- and post-pandemic periods.

## Data Availability

Non-identifiable individual-level serology data and aggregated surveillance data will be shared upon publication.

## Acknowledgements

We acknowledge the Seattle Children’s Hospital Microbiology Laboratory team, including Sean Murphy, MD, PhD and Drew Bell, PhD, for assisting in viral surveillance and specimen collection.

## Disclaimer

The findings and conclusions in this report are those of the authors and do not necessarily represent the official position of the US National Institutes of Health or Department of Health and Human Services.

