## Supplementary for "Multiplex serology reveals age-specific immunodynamics of endemic respiratory pathogens in the wake of the COVID-19 pandemic"

### Supplementary Material

I. Seattle serology: Number of study participants by pathogen and age group and p-values results for Kolmogorov-Smirnov tests by pathogen and age group comparing average antibody concentration levels in the 2020 baseline year to 2021 and 2022.

| Counts by pathogen and age group | | | |
|---|---|---|---|
| Age group | 2020 (N) | 2021 (N) | 2022 (N) |
| <1 | 40 | 42 | 19 |
| 1-2 | 70 | 78 | 61 |
| 3-4 | 47 | 69 | 57 |
| 5-10 | 166 | 196 | 154 |
| 18-49 | – | 128 | 99 |
| 50-64 | – | 147 | 134 |
| 65+ | – | 141 | 153 |

| Kolmogorov-Smirnov p-value results by pathogen and group comparing antibody concentration levels in 2021 to 2020 baseline. * denotes a significant difference between antibody concentration levels by year (p-value < .05) and ** denotes a significant difference after adjustment for multiple comparisons (p-value < .00125). | | | | |
|---|---|---|---|---|
| **Pathogen** | **<1 yo** | **1-2 yo** | **3-4 yo** | **5-10 yo** |
| CoV-HKU1 Spike | 0.1850 | 0.0006** | 0.3516 | 0.4177 |
| CoV-OC43 Spike | 0.3068 | 0.0013* | 0.0558 | 0.0644 |
| Flu B Brisbane | 0.1570 | 0.0101* | 0.4138 | 0.0744 |
| Flu-B-Phuket | 0.2657 | 0.0213* | 0.6430 | 0.0732 |
| Flu-H1-Michigan | 0.0292* | 0.0120* | 0.0587 | 0.5285 |
| Flu-H3-HongKong | 0.1007 | 0.0051* | 0.0554 | 0.0595 |

| | | | | |
|---|---|---|---|---|
| HCoV NL63 Spike | 0.0132* | 0.0015* | 0.0009** | 0.0050* |
| HCoV-229E Spike | 0.0130* | 0.0008** | 0.0013* | 0.1574 |
| RSV Pre-F | 0.2251 | 0.0007** | 0.0007* | 0.0035* |
| SARS-CoV-2 N | 0.3231 | 0.2574 | 0.3911 | 0.5299 |
| SARS-CoV-2 RBD | 0.2194 | 0.3259 | 0.2312 | 0.0586 |
| SARS-CoV-2 Spike | 0.1197 | 0.1934 | 0.2230 | 0.0256* |

Kolmogorov-Smirnov results by pathogen and group comparing concentration levels in 2022 to 2020 baseline. * denotes significant p-value < .05 and ** denotes significant p-value after adjustment for multiple comparisons. .00125

| **Pathogen** | **<1 yo** | **1-2 yo** | **3-4 yo** | **5-10 yo** |
|---|---|---|---|---|
| CoV-HKU1 Spike | 0.137 | 0.064 | 0.125 | 0.057 |
| CoV-OC43 Spike | 0.475 | 0.372 | 0.235 | 0.200 |
| Flu B Brisbane | 0.020* | 0.056 | 0.380 | 0.008* |
| Flu-B-Phuket | 0.043* | 0.107 | 0.834 | 0.002* |
| Flu-H1-Michigan | 0.058 | 0.010* | 0.045* | 0.067 |
| Flu-H3-HongKong | 0.048* | 0.143 | 0.001** | 0.040* |
| HCoV NL63 Spike | 0.252 | 0.302 | 0.043* | 0.609 |
| HCoV-229E Spike | 0.161 | 0.122 | 0.530 | 0.547 |
| RSV Pre-F | 0.034* | 0.114 | 0.004* | 0.317 |
| SARS-CoV-2 N | 0.003* | 0.000** | 0.000** | 0.000** |
| SARS-CoV-2 RBD | 0.000** | 0.000** | 0.000** | 0.000** |

| | | | | |
|---|---|---|---|---|
| SARS-CoV-2 Spike | 0.000** | 0.000** | 0.000** | 0.000** |

II. Antibody kinetics estimation based on Serosolver: Model estimates and sample size by age group, location, and pathogen. Models run with 5 chains and 700k iterations per chain.

| Study site, age group and pathogen | Antibody boost, 95% credible interval | Waning, 95% credible interval | N individuals (samples per person) |
|---|---|---|---|
| SOUTH AFRICA | | | |
| < 5 yo A/H3 | 5.50 (5.09, 5.87) | .13 (.08, .16) | 91 (2) |
| < 5 yo A/H1 | 5.86 (5.31, 6.40) | 0.09 (0.00, 0.15) | 90 (2) |
| < 5 yo B/Vic | 3.87 (3.12, 4.71) | .13 (.03, .21) | 90 (2) |
| < 5 yo B/Yam | 4.66 (4.17, 5.14) | .17 (.07, .25) | 90 (2) |
| 5-10 yo A/H3 | 2.90 (2.67, 3.13) | 0.08 (0.06, 0.09) | 285 (2) |
| 5-10 yo A/H1 | 2.88 (2.67, 3.09) | 0.04 (0.00, 0.07) | 285 (2) |
| 5-10 yo B/Vic | 2.14 (1.85, 2.45) | 0.04 (0.01, 0.07) | 285 (2) |
| 5-10 yo B/Yam | 3.17 (2.75, 3.83) | 0.09 (0.07, 0.11) | 285 (2) |
| 18 + A/H3 | 3.73 (3.52, 3.93) | 0.05 (0.04, 0.05) | 653 (2) |
| 18 + A/H1 | 3.62 (3.48, 3.80) | 0.05 (0.04, 0.06) | 653 (2) |

| | | | |
|---|---|---|---|
| 18+ B/Vic | 2.77 (2.56, 2.99) | 0.05 (0.05, 0.06) | 653 (2) |
| 18+ B/Yam | 3.13 (2.95, 3.31) | 0.06 (0.05, 0.07) | 653 (2) |
| KING COUNTY | | | |
| <5 yo A/H3 | 4.47 (3.84, 5.72) | .16 (.05, .27) | 396 (1) |
| <5 yo A/H1 | 4.11 (3.72, 4.57) | .21 (.13, .28) | 406 (1) |
| <5 yo B/Vic | 3.97 (3.42, 4.61) | .16 (.08, .24) | 407 (1) |
| <5 yo B/Yam | 4.47 (4.12, 4.84) | .25 (.18, .32) | 412 (1) |
| <5 yo RSV | 4.67 (4.29, 5.09) | 0.07 (0.02, 0.12) | 434 (1) |
| <5 yo CoV N | 4.77 (4.38, 5.17) | .15 (.01, .36) | 422 (1) |
| <5 yo CoV Spike | 4.97 (4.75, 5.21) | 0.05 (0.00, 0.10) | 429 (1) |
| <5 yo CoV RBD | 4.86 (4.60, 5.11) | 0.11 (0.05, 0.19) | 437 (1) |
| <5 yo HCoV 229E | 4.73 (3.94, 5.33) | 0.15 (0.09, 0,26) | 403 (1) |
| <5 yo HCoV NL63 | 4.09 (3.85, 4.38) | 0.03 (0.00, 0.08) | 397 (1) |
| <5 yo HCoV OC43 | 4.36 (3.01, 4.86) | 0.14 (0.09, 0.19) | 334 (1) |

| | | | |
|---|---|---|---|
| < 5 yo HCoV HKU1 | 4.06 (3.68, 4.46) | 0.06 (0.00, 0,13) | 419 (1) |
| 18+ yo A/H3 | 1.88 (1.53, 2.05) | 0.01 (0.00, 0.01) | 253 (2) |
| 18+ yo A/H1 | 1.90 (1.57, 2.08) | 0.07 (0.05, 0.08) | 254 (2) |
| 18+ yo B/Vic | 1.65 (1.62, 1.69) | 0.01 (0.01, 0.01) | 253 (2) |
| 18+ yo B/Yam | 1.36 (1.26, 1.55) | 0.03 (0.02, 0.06) | 254 (2) |
| 18+ yo RSV | 2.66 (2.50, 2.72) | 0.01 (0.01, 0.01) | 253 (2) |
| 18+ yo CoV N | 4.02 (3.84, 4.21) | 0.08 (0.01, 0.16) | 245 (2) |
| 18+ yo CoV Spike | 9.35 (8.95, 9.84) | 0.02 (0.00, 0.07) | 243 (2) |
| 18+ yo CoV RBD | 9.99 (9.34, 10.77) | 0.08 (0.00, 0.28) | 245 (2) |
| 18+ yo HCoV 229E | 1.62 (1.14, 2.50) | 0.01 (0.01, 0.02) | 245 (2) |
| 18+ yo HCoV NL63 | 1.25 (1.14, 1.42) | 0.03 (0.00. 0.09) | 244 (2) |
| 18+ yo HCoV OC43 | 1.23 (1.12, 1.73) | 0.01 (0.00, 0.04) | 243 (2) |
| 18+ yo HCoV HKU1 | 2.55 (2.40, 2.80) | 0.01 (0.01, 0.02) | 246 (2) |

III. Pairwise correlations in antibody concentration levels by pathogen and age group using the MSD multiplex assay, King County population. Each panel represents the correlation between antibody levels for two pathogens in the same individual at any of the sampled time points, grouped by age. Darker blue areas represent higher correlations. Note the higher correlations across SARS-CoV-2 antigens, alpha and beta coronavirus antigens, and influenza antigens.

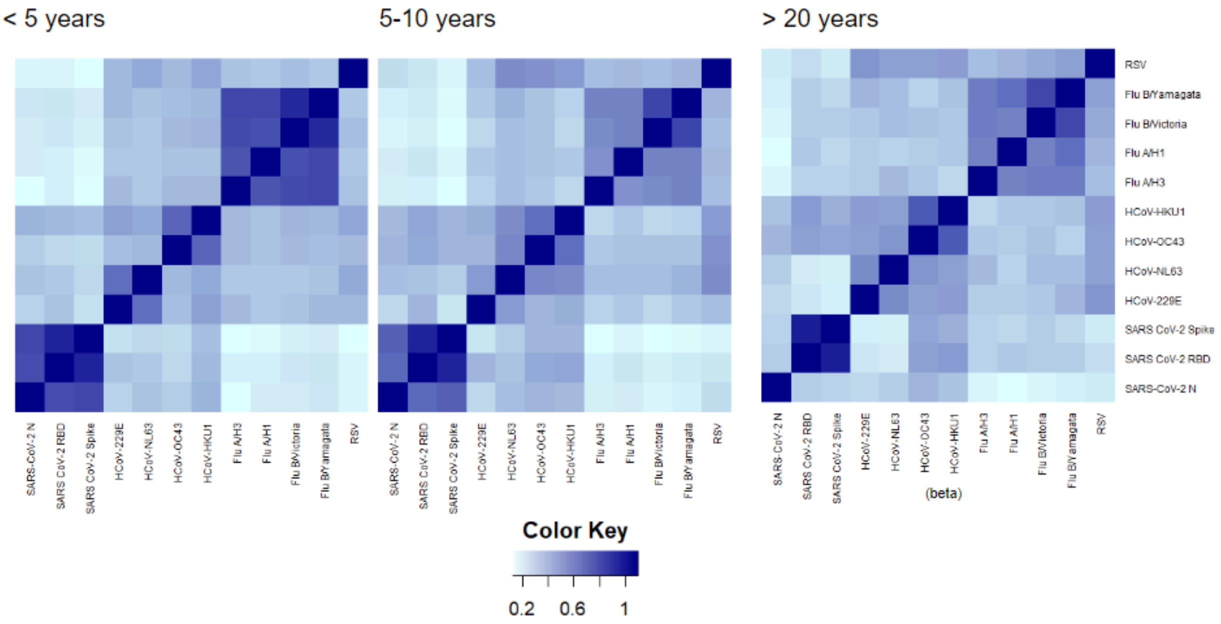


IV. Seasonal incidence of influenza-like Illness (ILI) in Washington State from 2016-2020 (top) and 2019-2023 (bottom), as reported by CDC surveillance.

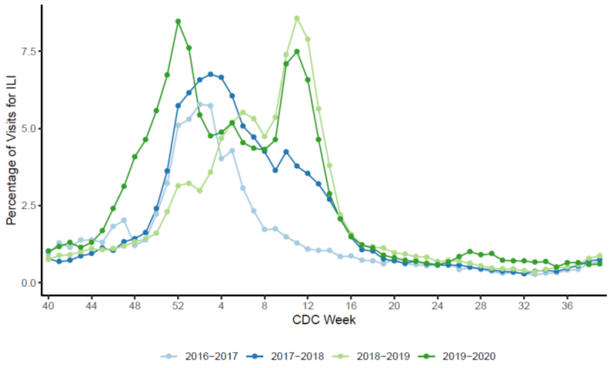

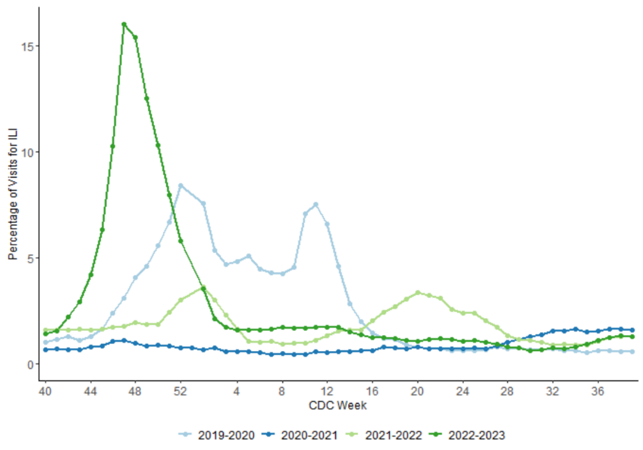


V. South Africa population attack rate observed in empirical data compared to beta distribution prior on attack rates used for Serosolver analyses.

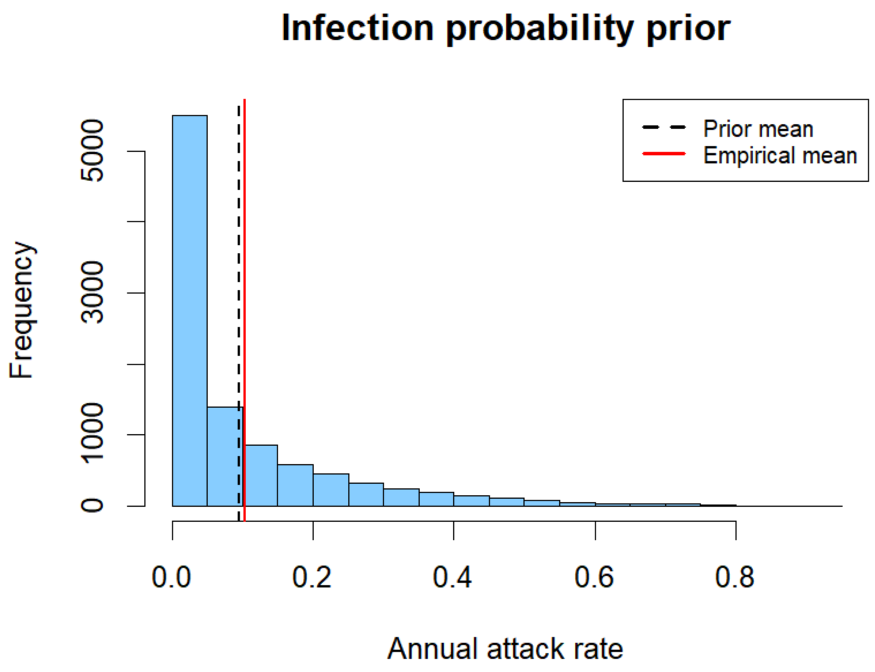

VI. Influenza transmission model parameter table

a. Uniform immunity structure (null model)

| Model Parameters | Definition | Source | Value |
|---|---|---|---|
| $1/v$ | Duration of immunity (years) | Fixed | 4 |
| $\beta_1$ | Amplitude of seasonal forcing | Fit | .28 |
| $1/\Upsilon$ | Infectious period (days) | Fixed | 2.3 |
| $1/\delta$ | Latent period (days) | Fixed | 2 |
| $\Phi$ | Phase shift | Fit | 4.59 |
| $\beta_0$ | Transmission coefficient | Fit | .53 |
| $\sigma_{1,\ 2,\ 3}$ | Proportion of reduced susceptibility via maternal antibodies in months 1,2,3 of infancy | Fixed | .08, .45, .45 |
| $h_{1,} h_{2\ ,} h_{3,,} h_{4,}$ | Age-specific hospitalization rates for 0-5, 5-10, 20-64, 65+ age groups | Fit | .12, .09, .06, .10 |
| $c_{1,} c_{2\ ,}$ | Reduced transmissibility via lockdowns March 2020-March 2021, April 2021-April 2022 | Fit | .57, .73 |

b. Tiered immunity structure model

| Model Parameters | Definition | Source | Value |
|---|---|---|---|
| $1/v$ | Duration of immunity (years) in individuals > 5 yrs | Fixed | 4 |
| $r$ | Change in duration of immunity in children < 5yrs, relative to older individuals (duration of immunity in children is r/$v$) | Fit | 0.49 |
| $\beta_1$ | Amplitude of seasonal forcing | Fit | .32 |

| | | | |
|---|---|---|---|
| $1/\Upsilon$ | Infectious period (days) | Fixed | 2.27 |
| $1/\delta$ | Latent period (days) | Fixed | 2 |
| $\Phi$ | Phase shift | Fit | 4.61 |
| $\beta_0$ | Transmission coefficient | Fit | .51 |
| $\sigma_{1,\ 2,\ 3}$ | Proportion of reduced susceptibility via maternal antibodies in months 1,2,3 of infancy | Fixed | .08, .45, .45 |
| $h_{1,}\ h_{2\ ,}\ h_{3,,}\ h_{4,}$ | Age-specific hospitalization rates for 0-5, 5-10, 20-64, 65+ age groups | Fit | .13, .20, .08, .08 |
| $c_{1,}\ c_{2\ ,}$ | Reduced transmissibility via lockdowns March 2020-March 2021, April 2021-April 2022 | Fit | .50, .61 |
